# The prevalence of adaptive immunity to COVID-19 and reinfection after recovery – a comprehensive systematic review and meta-analysis

**DOI:** 10.1101/2021.09.03.21263103

**Authors:** Tawanda Chivese, Joshua T. Matizanadzo, Omran A. H. Musa, George Hindy, Luis Furuya-Kanamori, Nazmul Islam, Rafal Al-Shebly, Rana Shalaby, Mohammad Habibullah, Talal Al-Marwani, Rizeq F Hourani, Ahmed D Nawaz, Mohammad Z Haider, Mohamed M. Emara, Farhan Cyprian, Suhail A. R. Doi

**Affiliations:** Department of Population Medicine, College of Medicine, QU Health, Qatar University, Doha, Qatar; Department of Public Health and Primary Care, Brighton and Sussex Medical School, United Kingdom; UQ Centre for Clinical Research, The University of Queensland, Herston, Australia; Department of Public Health, QU Health, Qatar University, Doha, Qatar; Immunology section, Basic Medical Sciences Department, College of Medicine, QU Health, Qatar University, Doha, Qatar; Microbiology section, Biomedical and Pharmaceutical Research Unit, QU Health, Qatar University, Doha, Qatar

**Keywords:** COVID-19, SARS-CoV-2, adaptive immunity, antibodies, reinfection, preventive efficacy

## Abstract

**Objectives:** This study aims to estimate the prevalence and longevity of detectable SARS-CoV-2 antibodies as well as memory cells T and B after recovery. In addition, the prevalence of COVID-19 reinfection, and the preventive efficacy of previous infection with SARS-CoV-2 were investigated.

**Methods and analyses:** A synthesis of existing research was conducted. The Cochrane Library for COVID-19 resources, the China Academic Journals Full Text Database, PubMed, and Scopus as well as preprint servers were searched for studies conducted between 1 January 2020 to 1 April 2021. We included studies with the relevant outcomes of interest. All included studies were assessed for methodological quality and pooled estimates of relevant outcomes were obtained in a meta-analysis using a bias adjusted synthesis method. Proportions were synthesized with the Freeman-Tukey double arcsine transformation and binary outcomes using the odds ratio (OR). Heterogeneity between included studies was assessed using the I2 and Cochran’s Q statistics and publication bias was assessed using Doi plots.

**Results:** Fifty-four studies, from 18 countries, with around 12 000 000 individuals, followed up to 8 months after recovery were included. At 6-8 months after recovery, the prevalence of SARS-CoV-2 specific immunological memory remained high; IgG – 90.4% (95%CI 72.2-99.9, I^2^=89.0%, 5 studies), CD4+ - 91.7% (95%CI 78.2 – 97.1, one study), and memory B cells 80.6% (95%CI 65.0-90.2, one study) and the pooled prevalence of reinfection was 0.2% (95%CI 0.0 – 0.7, I^2^ = 98.8, 9 studies). Individuals previously infected with SARS-CoV-2 had an 81% reduction in odds of a reinfection (OR 0.19, 95% CI 0.1 - 0.3, I^2^ = 90.5%, 5 studies).

**Conclusion:** Around 90% of people previously infected with SARS-CoV-2 had evidence of immunological memory to SARS-CoV-2, which was sustained for at least 6-8 months after recovery, and had a low risk of reinfection.

**Registration:** PROSPERO: CRD42020201234

**What is already known on this topic:** Individuals who recover from COVID-19 may have immunity against future infection but the proportion who develop immunity is uncertain. Further, there is uncertainty about the proportion of individuals who get reinfected with COVID-19.

**What this study adds:** Using data from 54 studies with follow up time up to 8 months after recovery, during the period February 2020-February 2021, we found that, post-COVID-19, up to 90% of individuals had antibodies and memory T and B cells against SARS-CoV-2. We also found a pooled prevalence of reinfection of 0.2%, and that infection conferred an 81% decrease in odds of reinfection with SARS-CoV-2, compared to unimmunized individuals without previous COVID-19.

This review of 12 million individuals presents evidence that most individuals who recover from COVID-19 develop immunological memory to SARS-CoV-2, which was still detectable for up to 8 months. Further, reinfection after recovery from COVID-19 was rare during the first 8 months after recovery, with a prevalence below 1%, while prior infection confers protection with an odds ratio of 0.19 and a preventive efficacy of 80% at a baseline prevalence of 5% for COVID-19 in a community.

**Implications of all the available evidence:** Individuals with a history of COVID-19 infection have immunity against the disease for up to 8 months, although this period could be longer. These individuals could be prioritized last for COVID-19 vaccinations or considered for single dose vaccinations.

**Strengths:** This comprehensive review addresses key questions on prevalent immunological memory and risk of reinfection in individuals with prior confirmed COVID-19 using robust systematic review methods.

**Limitations:** Some of the included studies which examined prevalent immunological memory were small studies which were affected by loss to follow up. The review did not examine evidence for immunity against the new divergent variants, which may be more likely to have immune evasion behaviour and may present a higher risk of reinfection. Lastly, the review did not examine the effect of the severity of COVID-19 on both immunological memory and the risk of reinfection.

## Introduction

Despite the availability of several efficacious and safe vaccines against COVID-19 (1, 2), access to the vaccines is limited, especially for low and middle-income countries (LMICs) (3). With the pandemic showing no signs of abating, a key question that remains unanswered is whether infection with COVID-19 confers immunity and how long that immunity lasts. If individuals with past COVID-19 infection have durable immunity, they may form a group that could be less prioritized for COVID-19 vaccination in resource-limited settings or subject to single-dose vaccination regimens in resource limited settings (4-6). However, the nature of, the protectiveness and duration of acquired immunity to COVID-19 is still not completely understood.

Several studies have shown that individuals infected with SARS-CoV-2 develop neutralizing antibodies (7, 8), and that, up to 8 months later, most individuals who recover from COVID-19 have evidence of humoral immunological memory (9-13). However, many of these studies involve small numbers of participants and suffer from loss to follow up. Therefore, it is still not clear what percentage of people with COVID-19 do have detectable antibodies against SARS-CoV-2 after recovery.

There is an increasing understanding of the role played by both cellular and humoral components of the adaptive immune response against SARS-CoV-2 infection (9, 14-17). Evidence suggests that even when there are no circulating antibodies, circulating memory T cells provide protection against clinical disease and death from infection by hepatitis B virus (18). This is likely to be true for SARS-CoV-2 which can progress relatively slowly to severe disease status, in a median of about 19 days (19), as this gives the cellular immune response the time it requires to muster. Findings from the few available studies suggest that it is likely that most individuals develop immunological memory to SARS-CoV-2 in the form of CD4+ and CD8+ T cells (9, 17). However, it is not yet clear what proportion of people who recover from COVID-19 have detectable cellular immunological memory to SARS-CoV-2 and for how long.

Measuring the proportions of individuals with evidence of immunological memory of SARS-CoV-2 gives a relatively good idea of immunity against the virus after recovery. An alternative way is through the measurement of the risk of reinfection after recovery from COVID-19. Initial findings from earlier studies suggested that reinfection with SARS-CoV-2 was not rare (20-23), and this was worsened by sensational news reporting. However, a growing understanding of COVID-19 has allayed these initial fears as most of early cases were more likely to be due to either prolonged viral shedding or reactivation of an incompletely cleared virus, that may have been harbored in the nasal cavity, termed repositivity (24-26). While repositive cases may not be as worrying as true reinfections, it is still important to understand how prevalent these cases are, and whether they are infective or not. Evidence from several case studies suggest that reinfection is still possible, with several confirmed reinfections having been reported so far (22, 23). Establishing reinfection prevalence is a difficult process in epidemiological studies as this requires viral sequencing from both the primary and secondary infection during large longitudinal cohorts (27), and not many studies can afford to do this on a large scale. Emerging data from a few large cohort studies (28-30) has shown that the prevalence of reinfection by SARS-CoV-2 lies anywhere between <1% and 5%. It is still not clear what proportion of individuals with COVID-19 get re-infected by the virus and if the protective immunity from previous infection by SARS-CoV-2 wanes over time. Longitudinal studies of the main seasonal coronaviruses; HCoV-NL63, HCoV-229E, HCoV-OC43 and HCoV-HKU1, have shown that acquired immunity is short lived and reinfection occurs more frequently after six to 12 months of recovery (31). This could be due to strain variation, which increases the risk of reinfection, a situation that may be similar to SARS-CoV-2 where divergent variants (32-34) have developed. Endemic human coronaviruses have seasonal outbreaks (35-38), but it is not yet clear whether SAR-CoV-2 will follow a similar pattern (39).

This research aims to estimate the prevalence of SARS-CoV-2 specific immunologic memory after recovery from COVID-19 and its efficacy in protecting against reinfection through synthesis of all existing research. Specifically, the research aims to estimate the prevalence of detectable SARS-CoV-2 specific IgG and IgA antibodies, memory CD4+, CD8+ and B cells after recovery, to estimate the prevalence of repositivity and reinfection after infection with SARS-CoV-2, and to estimate the protective efficacy of previous infection with SARS-CoV-2 against reinfection.

## Methods

### Study Design

The design and conduct of this systematic review and meta-analysis followed the Preferred Reporting Items for Systematic reviews and Meta-Analyses (PRISMA) guidelines (40). The protocol for this study is registered online on PROSPERO, the International prospective register of systematic reviews (CRD42020201234).

### Data sources and search methods for identification of studies

We searched for studies, without language restrictions, from 1 January 2020 till 1^st^ of April 2021, the Web of Science Clarivate, the China Academic Journals Full Text Database, PubMed, Scopus, and the databases of preprints (https://www.medrxiv.org/ and https://www.biorxiv.org/). All references of retrieved articles were manually screened for further studies. The search strategy is shown in Supplementary Doc 1.

### Procedure for selection of studies

Articles retrieved from the search were exported to Endnote X7 where duplicates were removed and then uploaded for the initial screening using title and abstract on the Rayyan systematic review management website (https://www.rayyan.ai/).

Due to the large number of records identified, records were subdivided into four groups and for each group, two investigators then screened titles, abstracts and if necessary full articles for inclusion. The full text of the records identified from screening using titles and abstracts were then screened for eligibility independently by the two investigators. Disagreements were resolved by an independent third author from another group.

### Criteria for considering studies for the review

#### Types of studies

This synthesis included observational studies which reported the prevalence of SARS-CoV-2 specific IgG, IgA, CD4+, CD8+ and memory B cells during and after recovery from COVID-19, the prevalence of repositivity and reinfection with SARS-CoV-2. The study designs included were case series of individuals, cross sectional studies and cohort studies. Studies were included if participants had COVID-19 confirmed using polymerase chain reaction (PCR) or molecular antigen tests. For humoral immunity, eligible studies should have assessed the presence of detectable SARS-CoV-2 specific IgG and IgA. For cellular immunity, studies were eligible if they used flow cytometry to measure proportions of individuals with SARS-CoV-2 specific CD4+, CD8+ and memory B cells. Studies were excluded if they were experimental studies, as they may not be representative of cases, if participants did not have confirmed COVID-19 by molecular tests, if they measured immunity components during the infection phase, if they were seroprevalence studies in general populations, if time points were not clear and if the study used commercial samples which were not linked to participants.

### Outcomes

For the humoral and cellular immune response, we assessed the prevalence of detectable SARS-CoV-2 specific IgG, IgA, CD4+, CD8+ and memory B cells after recovery from COVID-19. We also assessed the prevalence of reinfection and repositivity after recovery. Protective efficacy of prior infection with SARS-CoV-2 against reinfection was assessed by comparing the number of positive cases between individuals with prior COVID-19 and those without, within the same cohort.

### Key definitions

We defined the post infection period as at least 21 days after symptom onset or confirmation of COVID-19 and confirmation of two negative COVID-19 PCR 24 hour apart (27). A limitation of this definition is that individuals with long COVID could still be included, although it is estimated that this proportion is low at around 1.5% (41). We defined SARS-CoV-2 immunity prevalence as the proportion of individuals seropositive for any of IgG, IgA, CD4+, CD8+ memory B cells against any of; SARS-CoV-2 spike, spike receptor binding domain (RBD) and the nucleocapsid at the time point measured. Notably, recent studies document that while SARS-CoV-2 spike-specific serum IgA levels decline quickly after infection, local concentrations at mucosal surfaces persist longer and include dimeric isoforms with potent neutralizing capacity (42, 43) and thus serum IgA may not be the best indicator of mucosal protection. For studies that reported estimates at different timepoints, we extracted data from the furthest time point.

COVID-19 reinfection is difficult to establish and is strictly defined as phylogenetically distinct genomic sequences in the first and second episodes (27). It is becoming apparent that some recovered people may have a positive COVID-19 because of prolonged viral shedding. We defined repositivity as any positive PCR test within the first 3 months after a PCR negative tested recovery from COVID-19. Because very few population-based studies have been able to establish reinfection using genetic sequencing, and therefore distinguish reinfection from a chronic infection reservoir, we considered all participants who test positive on PCR for COVID-19 after being confirmed negative, or full clinical recovery with a negative COVID-19 test at least 3 months after recovery, according to the United States Centers for Disease Control criteria (27). One drawback is that this definition could possibly result in an under-estimation of reinfection rates.

The effect of prior infection with SARS-CoV-2 in protection against future infection was defined similar to the vaccination effect, by calculation of the relative risk reduction. However, the relative risk reduction was recomputed from the odds ratio (OR) using the Stata module *logittorisk* given that the synthesis needs to be done (44, 45) on the OR scale.

### Data extraction

From each included study, two reviewers extracted data on study characteristics such as study authors, country of study, study setting, timepoints measured, length of follow-up, gender distribution, and mean or median age of participants. Because of the high number of studies included and the difficulties in locating the data that were required for this synthesis, the studies were grouped into four groups and a third author was required to double check extracted data from each pair of reviewers. To estimate the prevalence of adaptive immune responses, repositivity and reinfection, we extracted data on numbers of individuals, out of the total with confirmed COVID-19, with circulating SARS-CoV-2 specific IgG, IgA, CD4+, CD8+, memory B cells, and numbers of individuals who had a positive RT-PCR after confirmed recovery from COVID-19 at least 3 months post their initial diagnosis. If a study reported data on multiple timepoints, we extracted data from the latest timepoints. For each study we also extracted data on the type of test used to detect IgG, CD4+, CD8+ and memory B cells and which SARS-CoV-2 antigen (spike protein, spike RBD or nucleocapsid) the antibodies or cells were specific to.

### Assessment of the quality of and risk of bias in included studies

Two investigators independently assessed the included articles for methodological quality using the updated MethodologicAl STandard for Epidemiological Research (MASTER) scale (analytical studies) (46) and the tool described by Hoy et al, (prevalence studies) (47). Any differences were resolved by discussion and a third investigator was consulted if they failed to reach consensus. The tool by Hoy et al (47), has ten items which assess external validity (items 1-4) and internal validity (items 5 - 9) since external validity is part of methodological quality for prevalence studies. The MASTER tool was used to assess the quality of analytical studies which assessed the effect of prior infection on the risk of reinfection with SARS-CoV-2. The MASTER scale has 36 items assessing seven quality domains which are; equal recruitment (items 1 - 4), equal retention (items 5 - 9), equal ascertainment (items 10 - 16), equal implementation (items 17 - 22), equal prognosis (items 23 - 28), sufficient analysis (items 29 - 31) and temporal precedence (items 32 - 36).

### Data synthesis

We used tables to show descriptive data of included studies. For the prevalence objectives, we re-calculated prevalence estimates from each study using the number of cases with detectable SARS-CoV-2 specific IgG, IgA, CD4+, CD8+, memory B cells and cases with positive PCR (reinfection) after recovery from COVID-19 that was at or later than 3 months post initial diagnosis with COVID-19. We also carried out subgroup analysis of prevalence in the post-recovery period using three periods of 0 - 2 months, 3 – 5 months and at least 6 months. We used the quality effects model (48) to pool prevalence from studies, as it maintains a correct coverage probability and a less mean squared error when compared to the random effects model, when there is heterogeneity across studies (49).

We used the Freeman-Tukey double arcsine transformation (50) to stabilize the variances in all prevalence data. To investigate the protective effect of prior COVID-19 on the risk of reinfection, we calculated unadjusted odds ratios from included studies Results of meta-analyses were presented in tables and forest plots.

Between study heterogeneity was investigated using the I^2^ statistic and Cochran’s Q p-values and exact p-values were presented. Heterogeneity was considered low (I^2^ below 50%), moderate (I^2^ between 50 - 75%) and high (I^2^ above 75%). Doi plots (39) were used to visually assess small study effects in lieu of funnel plots as they are more reliable and easier to interpret. The LFK index was used to quantify Doi plot asymmetry. We used the *metan* package in Stata IC version 15 software (51) for all analyses.

### Ethics and dissemination

This study utilized published data and did not require ethical approval.

### Patient and Public Involvement

It was not appropriate or possible to involve patients or the public in the design, or conduct, or reporting, or dissemination plans of our research

## Results

### Search results and characteristics of included studies

A total of 9 706 records were identified from database searches and a final 54 studies (8-12, 16, 17, 24-26, 30, 52-94) from 18 countries were included after exclusion of ineligible studies (Fig. 1). The total number of individuals from included studies were around 12 000 000 individuals. Most (n=23) of the studies were from China, eight were from the USA and the remaining from several other countries (Fig. 2). The follow-up times ranged from 2 weeks to 8 months after recovery from COVID-19, and the studies were carried out during the period February 2020-February 2021. Twenty-four of the studies were case series, 20 were cohort studies, eight were cross-sectional and the remaining two were case-control studies. Most of the studies (n=38) were hospital-based, 14 were carried out in the community and the remaining study was in health care workers only. Four (11, 16, 57, 69) of the included studies were preprints at the time of publication of this review. The characteristics of the included studies are shown in Supplementary Table 1.

**Fig. 1.**
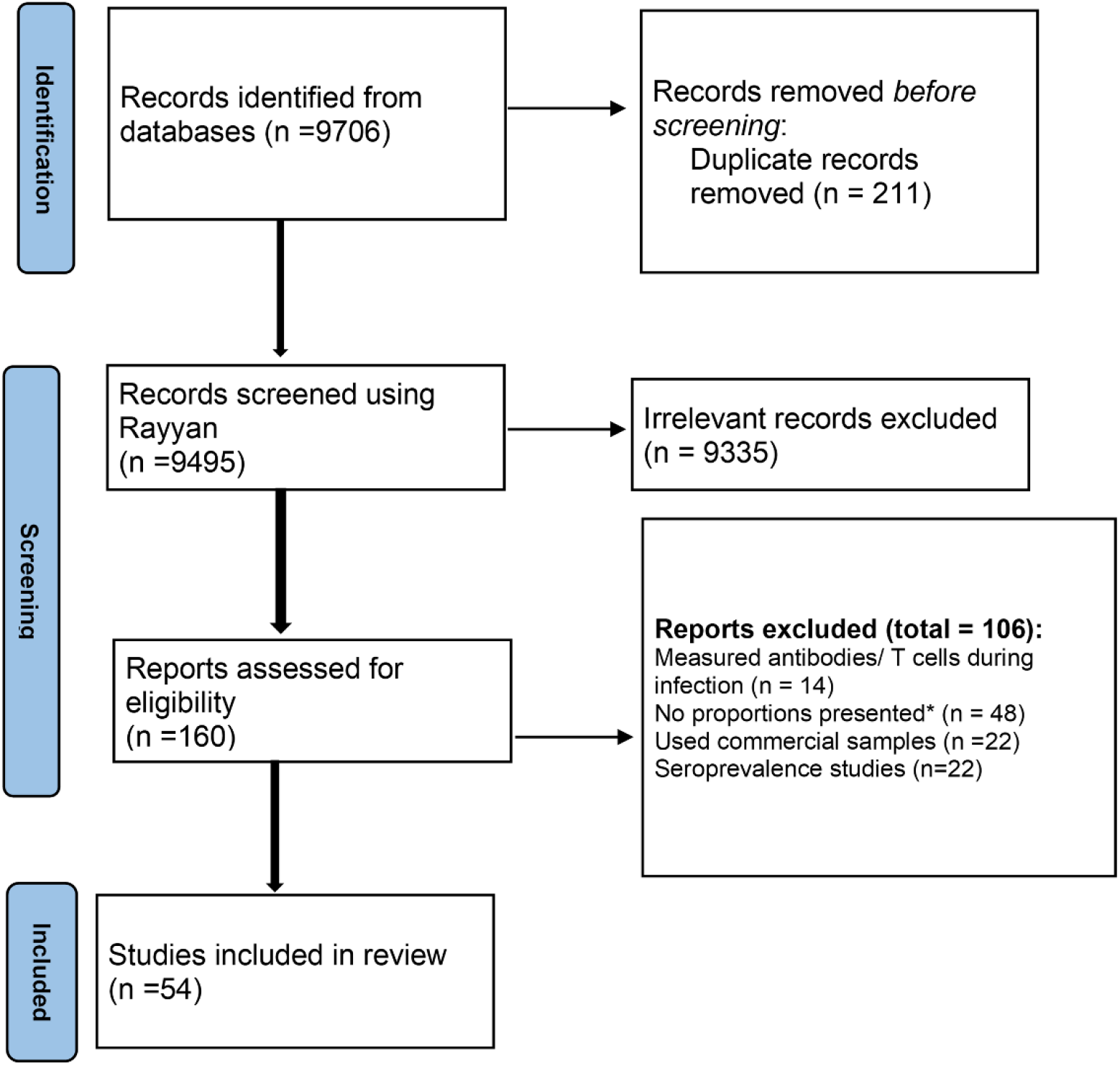
Flow chart showing the search and inclusion of studies

**Fig. 2.**
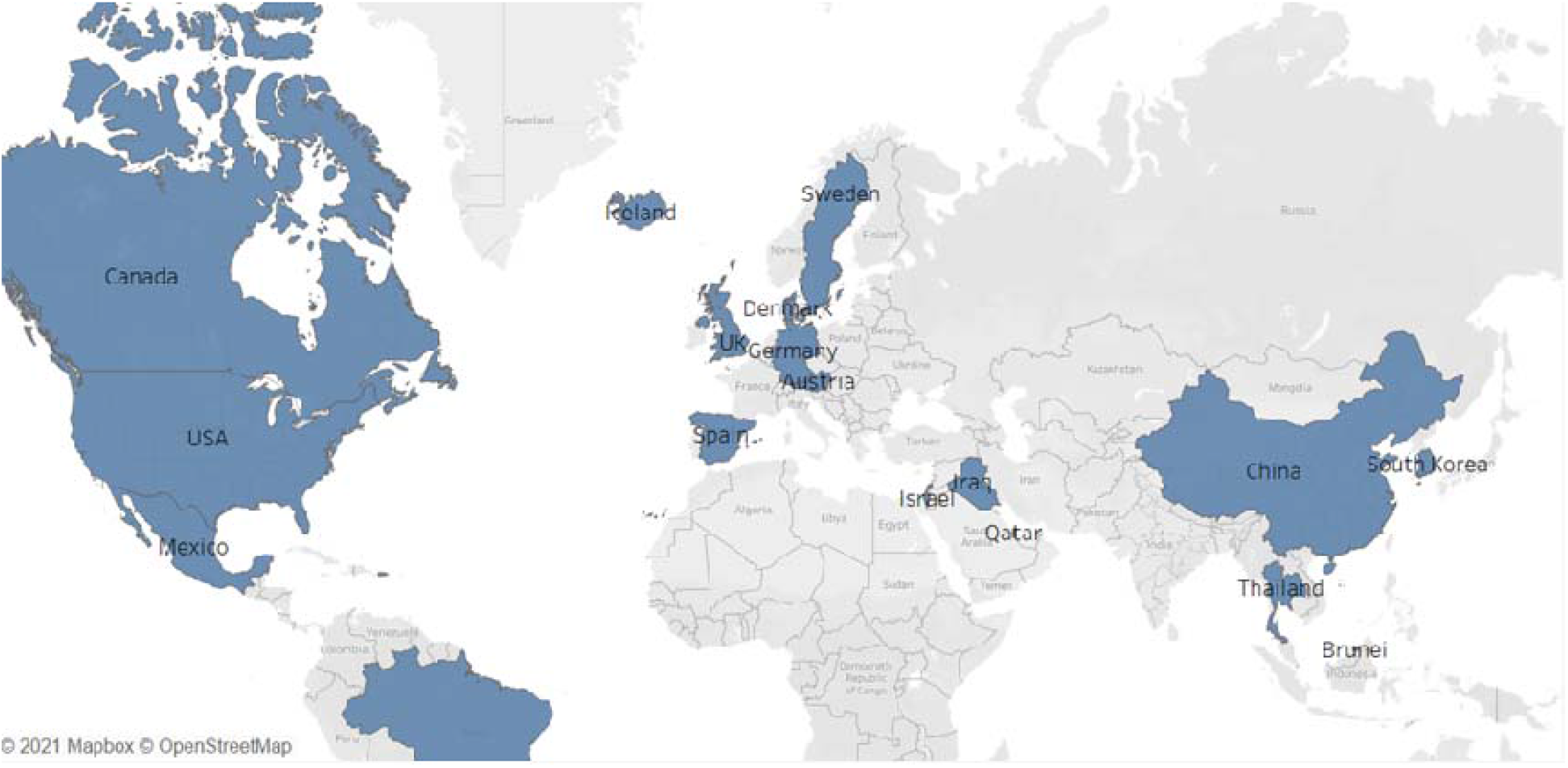
Map showing country of origin of included studies

### Assessment of internal validity and applicability

Using the Hoy risk of bias tool most of the studies (n=44) (8, 9, 12, 16, 17, 24-26, 30, 52, 54, 55, 58, 60, 61, 63-69, 71-75, 77-83, 87, 88, 90-95) were classified as moderate risk, 7 studies (11, 53, 56, 57, 62, 86, 89) as high risk and three studies (10, 60, 70) as low risk (Supplementary Table 2). The included studies had deficiencies in items related to external validity (Supplementary Table 2). The exceptions were a few studies where some forms of total sampling were employed. These included one Chinese study where the whole City of Wuhan was screened for COVID-19 after the first Chinese lockdown (10), an Austrian study where almost everyone in the country (70) was tested and a study of nearly four million people in Denmark (60). There were five studies (30, 54, 59, 70, 74) which compared new COVID-19 cases between individuals with prior infection and those without on reinfection/new infection which we assessed using the MASTER tool. The MASTER safeguard counts ranged from 14 to 21 out of a possible 36, with deficiencies in most of the studies, due to their observational nature, in the domains of equal recruitment, equal retention, equal ascertainment and equal prognosis (Supplementary Table 3).

### Prevalence of SARS-CoV-2 specific IgG and IgA antibodies after recovery from COVID-19

Data from 26 studies (8, 9, 11, 12, 16, 17, 56-59, 61, 62, 65, 68, 71, 73, 75, 77, 80, 83, 84, 86, 88, 90), with 3092 participants were available for evaluation of the prevalence of detectable IgG after recovery. The pooled prevalence of detectable IgG after recovery was 89.0% (95%CI 72.8 – 99.0, I^2^=89.7%, p<0.01, 8 studies) within 1 month, 92.6% (95%CI 86.9 – 96.9, I^2^=57.7%, p<0.01, 4 studies) at >1 - <3 months, 91.4% (95%CI 84.5-96.3, I^2^=84.3%, p<0.01, 9 studies) at 3-<6 months and 90.4% (95%CI 72.2-99.9, I^2^=89.0%, p<0.01, 5 studies) beyond 6 months (Fig 3 and Supplementary Figs. 1-4). There was no downward trend seen across these periods.

**Fig. 3.**
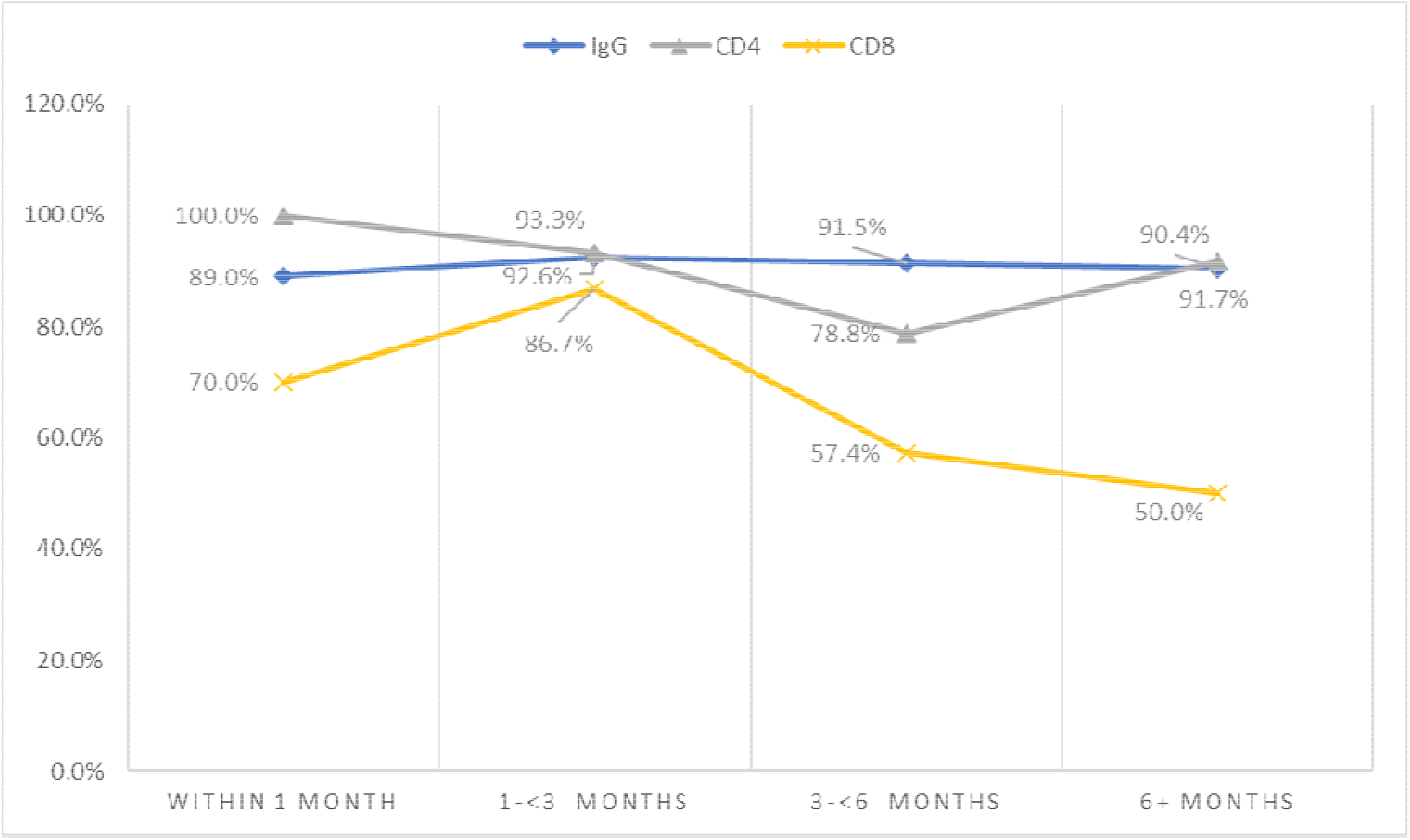
Prevalence of SARS-CoV-2 specific IgG, memory CD4+ and CD8+ cells after recovery from COVID-19

Only one study (69) reported data on the prevalence of SARS-CoV-2 specific IgA antibodies after recovery, with an estimate of 63.4% (95%CI 58.3 – 68.3) at 3 months after recovery.

### Prevalence of SARS-CoV-2 specific CD4+ and CD8+ after recovery from COVID-19

Data from four studies (9, 16, 17, 73), three from the USA and one from the UK, with a total of 118 participants resulted in a synthesized prevalence of detectable CD4+ T cells after recovery of 100% (95%CI 83.9 −100.0) within one month (17), 93.3% (95%CI 70.2 – 98.8) between 1 - 2 months USA (54), 78.8% (95%CI 65.1 – 88.0) at 4.5 months (16) and 91.7% (95%CI 78.2 – 97.1) at 6 - 8 months (7) (Fig 3 and Supplementary Fig. 5). Conversely, SARS-CoV-2 specific CD8+ T cells showed a steady decline after recovery from 70.0% (95%CI 48.1 – 85.5) within one month (17) to 50% (95%CI 34.5 – 65.5) at 6-8 months after recovery (7) (Fig 3 and Supplementary Fig. 6).

### Prevalence of SARS-CoV-2 specific memory B cells after recovery from COVID-19

Two studies (9, 67), both from the USA, reported data on the prevalence of SARS-CoV-2 specific memory B cells. In one study (67), most participants (prevalence 92.9%, 95% CI 68.5 – 98.7) had anti spike-RBD class switched memory B cells, between two to three months after recovery from COVID-19. The same pattern was observed in the other study (9), with 80.6% (95%CI 65.0 - 90.2) of the participants having RBD-specific memory B cells at 4-5 months.

### Reinfection after recovery from COVID-19

Nine studies, two from the UK (30, 54), and the remaining studies each from the USA (74), Austria (70), Denmark (60), Spain (96), Iraq (88), Qatar (87) and disputed territories (69), with a total of 257 448 participants, reported data on the prevalence of reinfection ≥3 after recovery from COVID-19. The reported prevalence ranged from 0.0% in Spain (52) to 5.7% in the USA (70) (Fig. 4). The pooled prevalence of reinfection was 0.2% (95% CI 0.0 – 0.7) with high heterogeneity (I^2^ = 98.8%, p<0.01) (Fig. 4). There was gross study asymmetry with smaller studies favouring more reinfection (Supplementary Fig. 7).

**Fig. 4.**
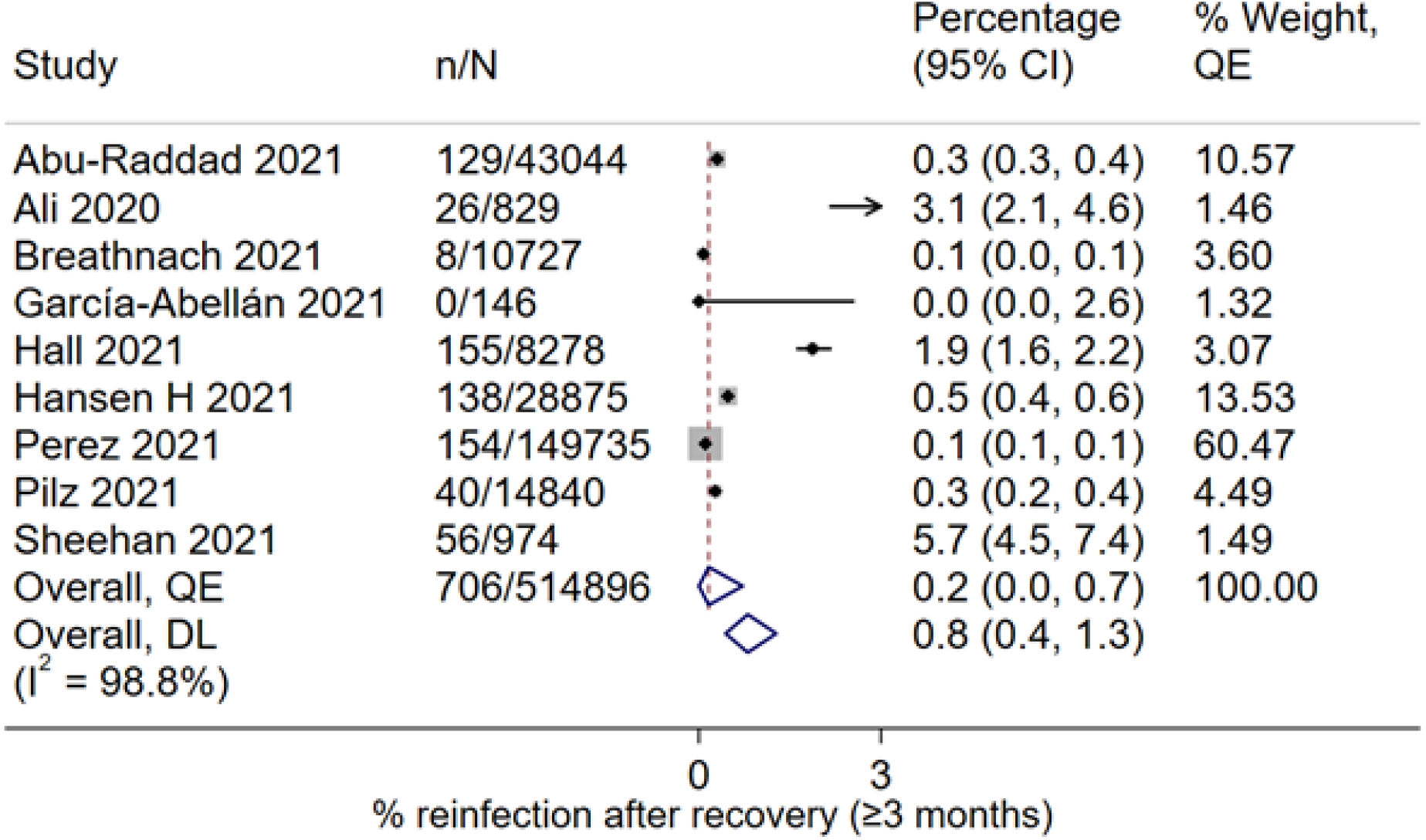
Prevalence of SARS-CoV-2 reinfection ≥3 months after recovery from COVID-19

In contrast to reinfection, the pooled prevalence of repositivity within one month was 0.9% (95%CI 0.0-8.0, I^2^ = 99.8%, p<0.01, 17 studies, Supplementary Fig 8). The pooled prevalence of COVID-19 repositivity at 2-3 months after recovery was 0.1% (95%CI 0.0 – 0.7, 7 studies), with substantial heterogeneity (I^2^ = 99.7%, p<0.01) and also gross study asymmetry with smaller studies favoring more repositivity (Supplementary Fig. 9).

### Comparison of risk of SARS-CoV-2 infection between individuals with and those without prior infection

Five studies with a total of 11 459 882 individuals compared infection with SARS-CoV-2 between individuals with a previous confirmed COVID-19 diagnosis and those who had no prior infection. Two of the studies were from the UK (30, 54), and one study each from Austria (70), Denmark (60) and the USA (74), and all the studies followed up participants for at least seven months. The odds ratio of infection by SARS-CoV-2 in individuals with prior COVID-19 compared to those without prior infection ranged from 0.06 in a study (76) from the UK to 0.41 in a study (80) from the USA (Table 1).

**Table 1.**
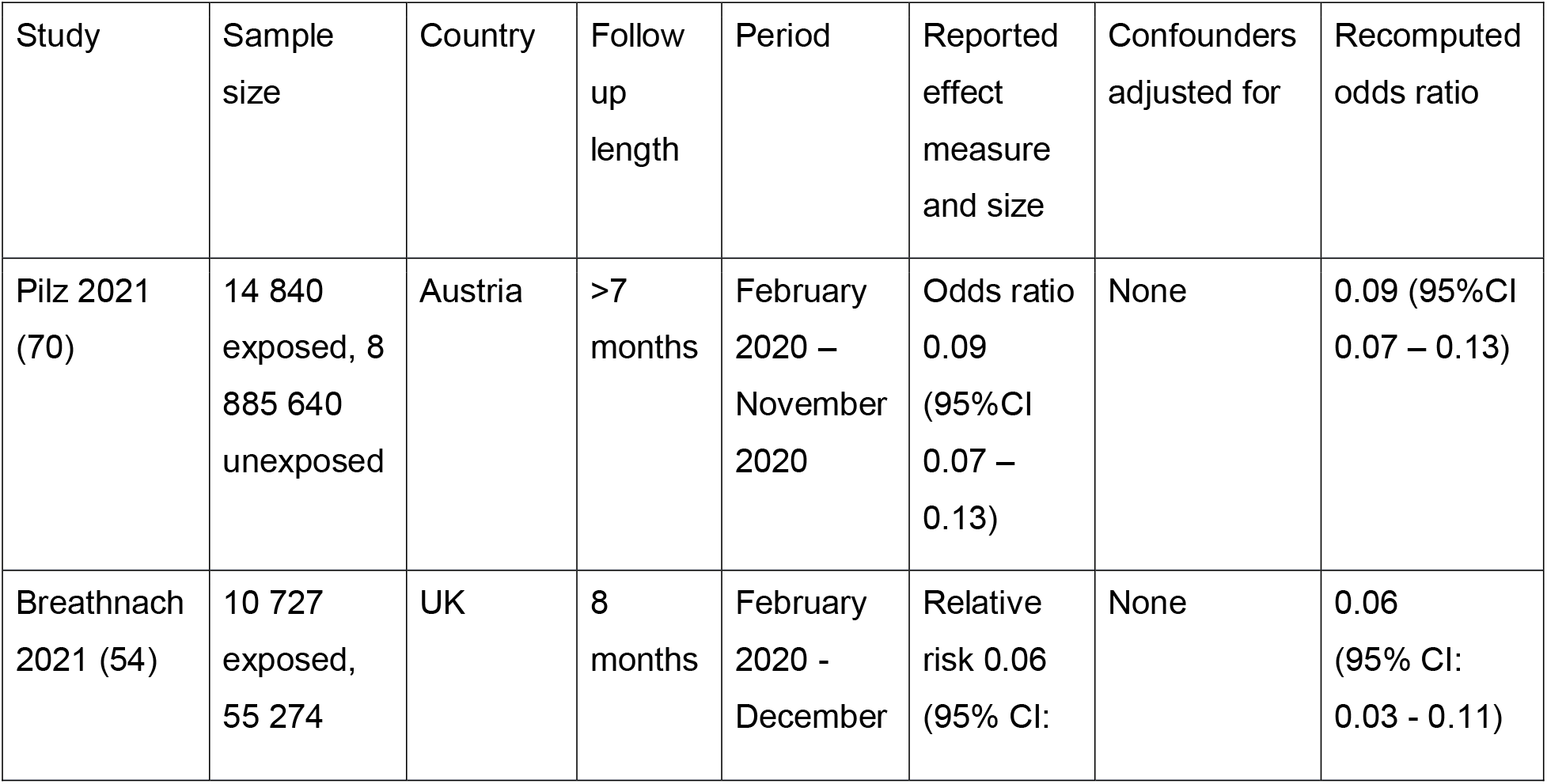
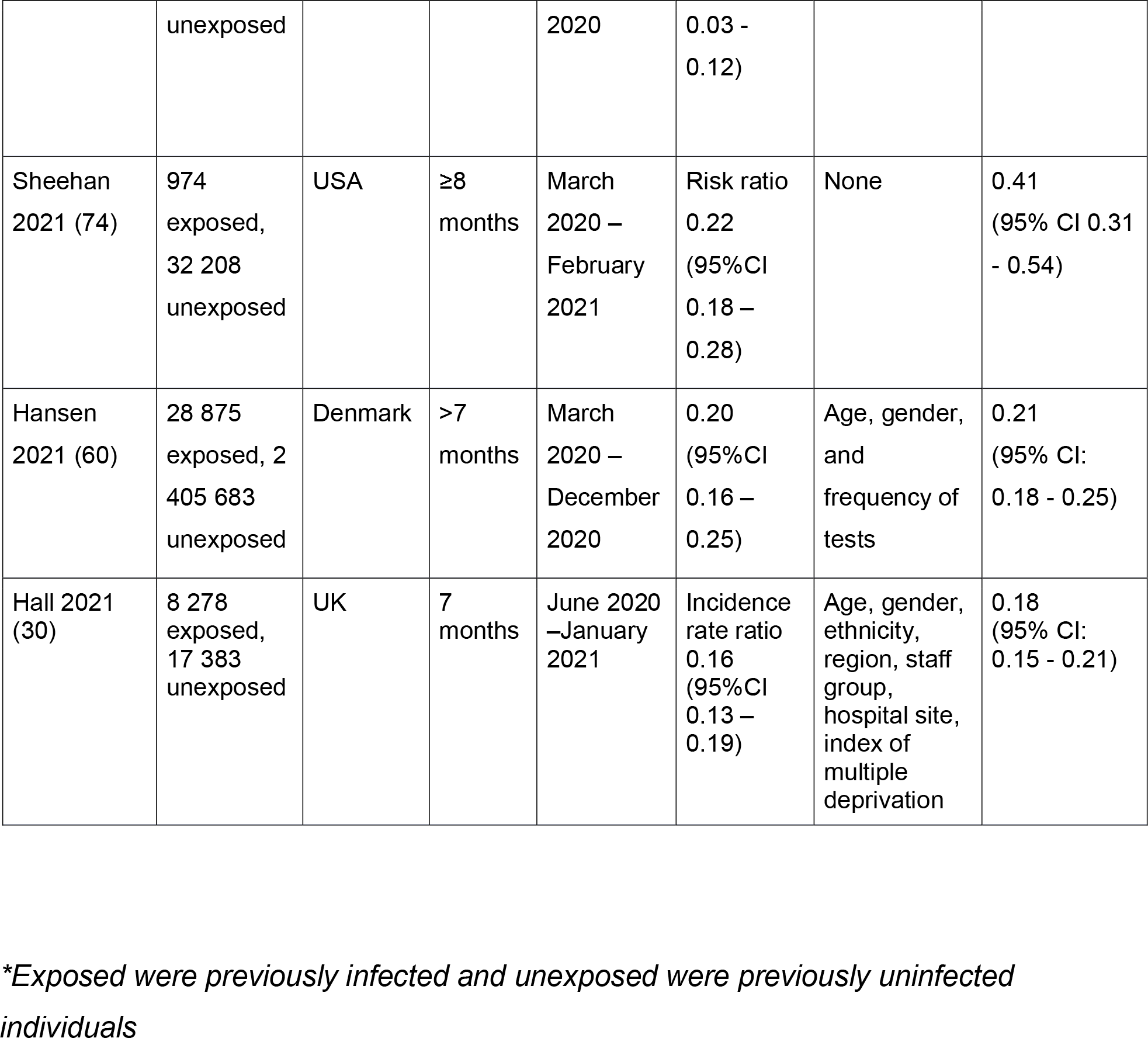
Studies comparing risk of infection by SARS-CoV-2 between previously infected and uninfected individuals.

In pooled analysis, the odds ratio for infection in individuals with compared to without prior COVID-19 was 0.19 (95%CI 0.11 – 0.32), with significant heterogeneity (I^2^ = 94.5, p<0.01) (Fig. 5). The studies were asymmetrical with smaller studies favoring less reinfection (Supplementary Fig. 10). Assuming a baseline risk of primary infection of 5%, this odds ratio translates (using logit to risk) to a relative risk reduction of 80.2% (95%CI 66.9% – 88.5%).

**Fig. 5.**
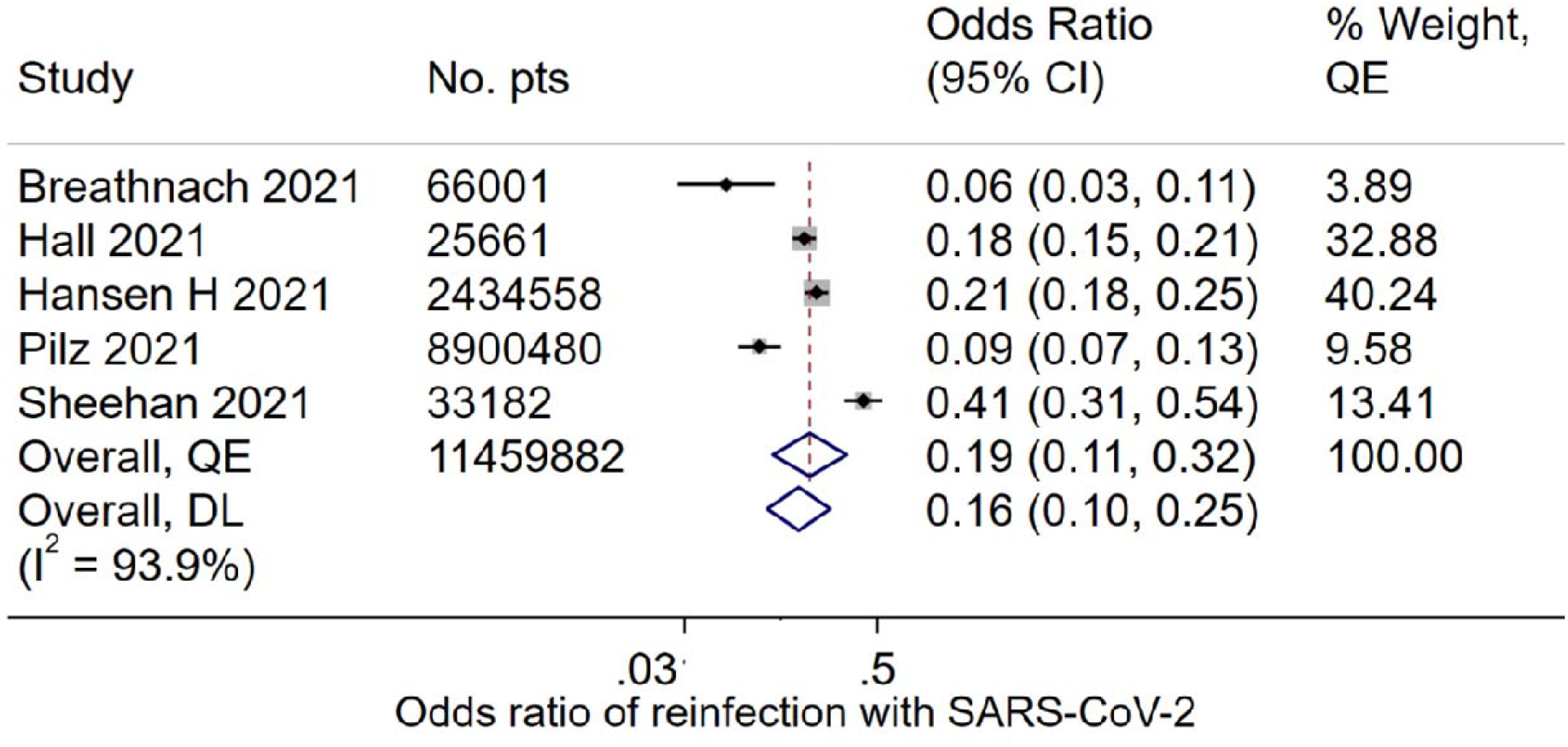
Pooled odds ratio of infection with SARS-CoV-2 in individuals with prior COVID-19 compared with those without prior infection

## Discussion

In this synthesis of 54 studies with follow-up times up to 8 months and carried out during the period February 2020-February 2021, we found a high prevalence of detectable SARS-CoV-2 specific immunological memory in the form of IgG antibodies, CD4+, and memory B cells in individuals who recovered from COVID-19. We found that the prevalence of reinfection after recovery from COVID-19 was very low and that prior infection with SARS-CoV-2 conferred an 80% protective efficacy against reinfection (assuming baseline prevalence of 5%). The existing reviews (23, 97-106) have not examined the question of prevalent immunity sufficiently (Supplementary Table 4).

The results of this study suggest that there is a sustained high prevalence of SARS-CoV-2 specific IgG antibodies, near 90%, up to 6-8 months after recovery from COVID-19, and data from one study showed a prevalence of IgA of 63% after recovery. Although we assessed the prevalence of detectable IgG, and not the actual antibody titres, research in primates has shown that even low circulating neutralizing antibody tires had a protective effect against COVID-19 (107, 108). This protective effect could be at the level of reducing severe COVID-19 and death from COVID-19, rather than stopping the infection in the upper respiratory tract. Sterilizing immunity that stops individuals from acquiring infection requires high antibody titres (9), however the level of antibody titres that provides sterilizing immunity is still not known. Therefore, it is still theoretically possible that individuals with detectable IgG may be infected by and are able to spread SARS-CoV-2. Longitudinal studies are needed to investigate the relationship between antibody titres and the risk of reinfection. Notably, it appears that the SARS-CoV-2 antibody titres are stable after recovery with a half-life of almost 5 months for spike IgG (9). Further, several studies (5, 6, 109, 110) have shown that, regardless of either antibody titre or the presence of detectable IgG in individuals with prior infection, the levels of IgG and neutralizing antibodies after a first dose vaccination, reached titres similar to those of SARS-CoV-2 naïve individuals after a second dose, and, in one study, more than 100 times those of naïve individuals (5). The findings from this current synthesis add to this body of knowledge and support single-dose vaccination for individuals with prior infection.

This synthesis suggests, for a period of at least 6-8 months after recovery, around 90% of individuals have evidence of SARS-CoV-2 specific memory B and memory CD4+ cells while about half have evidence of CD8+ cells. While the role of T cells in sterilizing immunity is thought to be limited, they are highly associated with ensuring less severe COVID-19 (73, 111). A diminished prevalence of cytotoxic CD8+ cells may imply that viral clearance is delayed in some individuals, in the event of reinfection. However, there is evidence of sustained high prevalence of T follicular helper cells (TFH) (9), a subset of CD4+ T cells that are the most important in helping memory B cells and in the production of neutralizing antibodies and long-term humoral immunity (99). A high prevalence of memory B cells at ≥6 months also suggests that immunological memory may be long lasting, at least to the time points measured in the included studies.

This synthesis suggests that although repeat test-positives are likely to occur in about 2% of individuals within 1 month of recovery, the prevalence of reinfection with SARS-CoV-2 is low, with only 0.2% reinfected during a period of up to 8 months after recovery. Further, prior infection with SARS-CoV-2 provides protection against reinfection with an efficacy of 80%, during the same period. While the presence of antibodies and memory T and B cells are evidence of immunological memory, prevalent reinfection is a stronger measure of clinical protection of prior infection (70). In a letter to the editor published at the time of finalization of this review, the prevalence of reinfection in a period of up to 12 months was 0.3% in Italy (112). Findings from this Italian study (101) also suggested a strong protective efficacy of prior infection with a hazard ratio of 0.06 (95%CI 0.005 – 0.08). In the USA, another study of 9119 individuals (113) with serial tests at least 90 days apart, during December 2019 to November 2020, published at the time of finalization of this review, also showed a low prevalence of reinfection of 0.7% (95%CI 0.5-0.9) (89). More research is also needed to measure protective efficacy in the long term.

A key consideration is that most of the studies in this synthesis included individuals infected with the original variant of SARS-CoV-2 and had follow up periods during the year 2020, which may not have covered exposure to new variants. The substantial changes to the SARS-CoV-2 spike protein and the RBD domain in the new variants (32-34), with the delta variant currently being dominant globally (114), may adversely affect immunological memory and increase the risk of reinfection. For example, high case counts and hospitalizations were observed in Manaus, Brazil, 7 months after seroprevalence data suggested that three-quarters of the population were previously infected with SARS-CoV-2 (115), suggesting that a large proportion of the population was still susceptible to infection by SARS-CoV-2 (116). While it is likely that the seroprevalence study overestimated the proportion infected with SARS-CoV-2, it is also equally likely that the resurgence in Manaus could have been driven by the emergence of the highly divergent and transmissible gamma (P1) variant which was first reported from Manaus (32, 116). Divergent lineages of SARS-CoV-2 are more likely to be associated with antigenic escape (116) and therefore result in higher chances of reinfection. Estimates of reinfection with divergent variants such as delta and Omicron are therefore unknown, and more research is required.

Our research has several limitations, one of which is that these findings cannot be extended beyond the time of follow up of the included studies. Another limitation is the heterogeneity in the studies that we included which was not reduced by subgroup analyses in measurements methods and follow up times. Many of the longitudinal studies in this meta-analysis suffered from loss to follow up and this may have affected their findings. Further, although we included a comprehensive number of studies, studies on the cellular immune response are lacking and the ones we included had very small sample sizes, implying that our estimates of this aspect of the adaptive immune response may change with the accumulation of more data. Many of the included studies were small observational studies which are easily affected by confounding. Lastly, the review did not examine evidence for immunity against the new variants and did not include studies with longer follow up.

## Conclusion

This synthesis shows around 90% of individuals have evidence of SARS-CoV-2 specific immunological memory. Further, the risk of reinfection is rare and in the first 6-8 months after infection, risk of re-infection decreased by 80%, though that could diminish after 8 months..

## Supporting information

Supplementary Document S1. Search Strategy

## Data Availability

All data are from published studies

## CRediT Authors’ contributions

TC - Conceptualisation, data curation, formal analysis, investigation, methodology, validation, resources, software, visualisation, writing – original draft, and writing – review & editing

JTM - Data curation, investigation, formal analysis, visualization, writing – review & editing

OAHM - Data curation, investigation, formal analysis, methodology, writing – review & editing

LFK - Investigation, methodology, formal analysis, writing – review & editing

NI – Data curation, investigation, writing – review & editing

RA – Data curation, investigation, formal analysis, writing – review & editing

RS – Data curation, investigation, formal analysis, writing – review & editing

MH – Data curation, investigation, formal analysis, writing – review & editing

TA – Data curation, investigation, formal analysis, writing – review & editing

RFH – Data curation, investigation, formal analysis, writing – review & editing

ADN – Data curation, investigation, formal analysis, writing – review & editing

MZH – Data curation, investigation, formal analysis, writing – review & editing

MME – Conceptualisation, investigation, writing – review & editing

FC – Conceptualisation, investigation, writing – review & editing

SARD - Conceptualisation, formal analysis, investigation, methodology, resources, software, supervision, validation, visualisation, writing – review & editing All authors critically reviewed the final manuscript.

TC and SARD had full access to and verified the data

