## Supplementary Document S1. Search Strategy for "The prevalence of adaptive immunity to COVID-19 and reinfection after recovery – a comprehensive systematic review and meta-analysis"

### **Supplementary Tables and Figures.**

**Supplementary Doc 1 - Search strategy**

Pubmed, Scopus and Web of Science Clarivate (Advanced search) (adapted for other database)

#1. ( immune OR immuned OR immunes OR immunisation OR vaccination OR vaccination OR immunization OR immunization OR immunisations OR immunizations OR immunise OR immunised OR immuniser OR immunisers OR immunising OR immunities OR immunity OR immunity OR immunizations OR immunize OR immunized OR immunizer OR immunizers OR immunizes OR immunizing OR antibodies OR antibodies OR antibodies OR antibody OR antibody OR immunoglobulins OR immunoglobulins OR antibody OR IgG OR IgM OR “immunoglobulin G” OR “immunoglobulin M” OR “immunoglobulin A” OR IgA OR T cells OR B cells OR “immune memory” OR “immunological memory” OR “memory cells” OR CD4 OR CD8 OR “memory B cells” OR “memory T cells” OR “humoral immunity” OR “cellular immunity” OR “adaptive immunity” OR “memory CD4 T cells” OR “memory CD8 T cells” OR “memory CD4+ T cells” OR “memory CD8+ T cells” OR “memory CD4^+^ T cells” OR “memory CD8^+^ T cells” OR reinfect* OR repositive)

#2. (COVID  OR  COVID19  OR  "SARS‐CoV‐2" OR "SARS‐CoV2" OR SARSCoV2 OR"SARSCoV‐2" OR "SARS coronavirus 2" OR "2019 nCoV" OR "2019nCoV" OR "2019‐novel CoV" OR "nCov 2019" OR "nCov 19" OR  "severe acute respiratory syndrome coronavirus 2" OR "novel coronavirus disease" OR "novel corona virus disease" OR "corona virus disease 2019" OR "coronavirus disease 2019" OR "novel coronavirus pneumonia" OR "novel corona virus pneumonia" OR "severe acute respiratory syndrome coronavirus 2") OR AB=(COVID  OR  COVID19  OR  "SARS‐CoV‐2" OR "SARS‐CoV2" OR SARSCoV2 OR"SARSCoV‐2" OR "SARS coronavirus 2" OR "2019 nCoV" OR "2019nCoV" OR "2019‐novel CoV" OR "nCov 2019" OR "nCov 19" OR  "severe acute respiratory syndrome coronavirus 2" OR "novel coronavirus disease" OR "novel corona virus disease" OR "corona virus disease 2019" OR "coronavirus disease 2019" OR "novel coronavirus pneumonia" OR "novel corona virus pneumonia" OR "severe acute respiratory syndrome coronavirus 2")

#3. #1 AND #2
Filters

Time=2020‐2021

### **Supplementary Table 1 – Characteristics of included studies**

| **Citation (First Author year)** | **Country** | **Setting hospital or community based** | **Study design** | **Total sample size** | **Age** | **Gender** | **Severity** | **Timepoints measured** | **domain** |
| --- | --- | --- | --- | --- | --- | --- | --- | --- | --- |
| Zhu 2020 (1) | China | Hospital | Cohort | 98 | 52 years (IQR, 37.8 ‐ 59) | 67.3% female | Hospitalized and discharged | Median 21 (IQR 17-28) | Reinfection |
| Zheng 2020 (2) | China | Hospital | Cohort | 20 | Age range 23-57 | 70% male | Hospitalized and discharged | 2 weeks after recovery | Re-infection |
| Zhao Y 2020 (3) | China | Hospital | Cohort | 55 | Mean 47.74 years | 41.82% female | Hospitalized and discharged | 3 months after recovery | Humoral |
| Zhao J 2020 (4) | China | Hospital | Cohort | 173 | Median 48 years (IQR, 35–61 years) | 49% male | Hospitalized and discharged | 39 days from disease onset | Humoral, Post humoral |
| Zhang 2020 (5) | China | Hospital | Case series | 127 | Mean 44.2 | 50% female | NR | 0 - 38 days after recovery | Humoral |
| Yuan B 2020 (6) | China | Hospital | Cohort | 182 | Mean 46.4 ± 17.1 years | 46.2% males | Hospitalized and discharged | Day 14 after recovery | Re-infection |
| Yuan J 2020 (7) | China | Hospital | Cross sectional | 172 | Median 28 yrs | 61% female | Hospitalized and discharged | 2 weeks after recovery | Re-infection |
| Ye 2020 (8) | China | Hospital | Cohort | 55 | Median 37 (range 22 - 67) | 65.45% female | Hospitalized and discharged | 4-17 days after recovery | Re-infection |
| Xiao 2020 (9) | China | Hospital | Cross sectional | 70 | Mean 57 yrs | 44.3% male | Moderate | Median 22 days after recovery | Reinfection |
| Xiang 2020 (10) | China | Hospital | Case series | 85 | Range 32 – 65 years | 63.5% females | Both asymptomatic and symptomatic | ≥30 days PSO | Humoral |
| Wu X 2021 (11) | China | Community | Case series | 20280 | Median 56 years | 46.51% males | Asymptomatic | 8 months | Re-infection |
| Wu J 2020 (12) | China | Hospital | Cross sectional | 60 | Median 46.5 (IQR 33.5-58.5) | 43.3% female | Hospitalized and discharged | Median 21 days PSO | Re-infection |
| Wong 2020 (13) | Brunei | Hospital | Cross sectional | 106 | Median age 47 | 60.3% males | Both asymptomatic and symptomatic | Median 32days (IQR 28.75 - 33.5) PSO | Re-infection |
| Shu 2020 (14) | China | Hospital | Case series | 131 | Mean 51.4 years | Male - 68.7% | Hospitalized and discharged | 21-40 days PSO | Humoral |
| Sheehan 2021 (15) | USA | Community | Cohort | 33182 | Mean 51.7 yrs (SD 22.2) | 54.7% female | Both asymptomatic and symptomatic | >9 months | Reinfection and effectiveness |
| Rydyznski 2020 (16) | USA | Community | Case series | 54 | Median 55.5 | 71% Male | Severely ill | 3 weeks post blood for convalescent (4 - 37 days) | Cellular and humoral |
| Qiao 2020 (17) | China | Hospital | Cohort | 15 | Mean 36.7 years (± 14.91) | 53.3% Male | Hospitalized and discharged | 2nd and 4th weeks after discharge | Reinfection |
| Prévost 2020 (18) | Canada | Community | Cross sectional | 98 | Mean 55 years | 50% male | Both asymptomatic and symptomatic | 8 - 14days PSO | Post Humoral |
| Pilz 2021 (19) | Austria | Hospital | Cohort | 8900480 | Median 39.8 yrs | 62.5% female | Both asymptomatic and symptomatic | 7 months | Reinfection |
| Perez 2021* (20) | Israel | Hospital | Case series | 149735 | Mean 31.5 (SD± 19.7) | 61% male | Symptomatic  Hospitalized and discharged | 3 months | Reinfection |
| Peng 2021 (21) | China | Community | Cohort | 20 | Median 51.5 years (range 45–65) | 55% Male | Hospitalized and discharged | 5 (range 5 – 33 days) and 230 (range 221 – 248 days) days after symptom onset | Post humoral |
| Olea 2021 (22) | Spain | Hospital | Case series | 35 | Median age 62.5 | 66% Male | Hospitalized and discharged | 4 months | Post humoral |
| Ogega 2020 (23) | USA | Hospitalized | Case series | 14 | Mean 57.6 years | 57% male | Hospitalized and discharged | 54 days after symptoms | Cellular immunity |
| Murillo-Zamora 2021 (24) | Mexico | Community | Cohort | 100432 | 81.8% were ≤49 | 53.9% female | NR | 56 days (IQR 40-81) | Reinfection |
| Liu 2020 (25) | China | Community | Case series | 150 | Mean 51.5 yrs | 49.3% male | Hospitalized and discharged | 38-39 days from symptom onset | Re-infection |
| Li Y 2020 (26) | China | Hospital | Cohort | 13 | Mean age 52.8 ± 20.2 years | 46.1% male | Hospitalized and discharged | Median 32.5 (IQR 30.5-39.25) | Reinfection |
| Ko 2020 (27) | South Korea | Hospital | Cross sectional | 64 | Median 32 years | 44% male | Asymptomatic,  Mild | 43 days after symptom onset | Post humoral |
| Jiang 2020 (28) | China | Hospital | Cross sectional | 35 | Median 45.2 (IQR 30-56) | 100% female | Both asymptomatic and symptomatic | Median 32.5 (IQR 31.25-36) | Reinfection |
| Jia 2020 (29) | China | Hospital | Case series | 19 | Mean 9 years (7months-13yrs) | 47.3% male | Hospitalized and discharged | 11-27 days PSO | Humoral, Post humoral |
| Huynh 2021 (30) | USA | Hospital | Cohort | 153 | Mean 49 years (18 – 82) | 62.1% female | Hospitalized and discharged | 7-211 days post-symptom onset | Humoral |
| Huang 2020 (31) | China | Hospital | Cohort | 414 | Range 0-86 yrs | 47.1% male | Hospitalized and discharged | Every 3 to 5 days | Reinfection |
| Havervall 2020* (32) | Sweden | Hospital | Case series | 59 | Mean 44 years (SD± 12) | 69% male | Hospitalized and discharged | 4 months | Post humoral |
| Hansen 2021 (33) | Denmark | Community | Cohort | 2434558 | NR | NR | NR | >7 months | Re-infection and effectiveness |
| Hansen 2021 (34) | Denmark | Hospital | Case series | 350 | Median 52 (41-63 years) | 43.1% male | Both asymptomatic and symptomatic | Convalescent 4-11 weeks after symptoms onset | Post humoral |
| Hamed 2020 (35) | Qatar | Hospital | Case series | 63444 | Recurrent positive median: 37.3yrs (IQR 11-74) | 79% male | Both asymptomatic and symptomatic | mean 29 days, range 21-84 after recovery | Reinfection |
| Hall 2021 (36) | UK | Healthcare workers | Cohort | 25661 | Median 46yrs (IQR 18.6-78.4) | 82.4% female | Both asymptomatic and symptomatic | ≥7 months | Reinfection and effectiveness |
| Grifoni 2021 (37) | USA | Hospital | Case-control | 40 | Cases (20–64 (median = 44, IQR = 9)), Controls (20–66 (median = 31, IQR = 21)) | 40% male | Recovered, symptomatic,  Non-hospitalized | 20 - 35 days PSO | Cellular immunity |
| Gudbjartsson 2020 (38) | Iceland | Community | Case series | 487 | nr | nr | Hospitalized and discharged | 4 months | Humoral |
| García-Abellán 2021 (39) | Spain | Hospital | Case series | 116 | Median 64 yrs | 60% male | Hospitalized and discharged | 6 months | Reinfection, Post humoral |
| Fischer 2021 (40) | Germany | Hospital | Case series | 41 | Mean 54 years (± 8.4) | 57% Male | Mild to moderate | Convalescent 28 - 288 days after recovery | Post humoral and cellular immunity |
| Fendler 2020* (41) | UK | Hospital | Case-Control | 144 | Median 59.4 years | 49% Male | Hospitalized and discharged | 8 - 202 days after recovery | Cellular immunity |
| De Giorgi 2021* (42) | USA | Community | Case series | 202 | Mean 47 years (19-79) | 45% Males | Asymptomatic (3%), moderate 90% | 5 months | Post humoral |
| Dan 2021 | USA | Hospital | Case series | 188 | Median 40 years | 43% male | 93% mild – never hospitalized | 8 months | Cellular, Humoral |
| Chirathaworn 2020 (43) | Thailand | Hospital | Case series | 217 | Median 33yrs (IQR 25–47) | Male 42.4%; Female 57.6% | Both asymptomatic and symptomatic | Days 28 – 142 PSO | Reinfection, Post humoral |
| Chen 2020 (44) | China | Hospital | Cohort | 1067 | Median 60 years (IQR 49-69) | 41.6% male | Hospitalized and discharged | 50 Days (IQR 36.5 - 59.5) PSO | Reinfection |
| Cao S 2020 (45) | China | Population-based census | Cross sectional | 34420 | NR | 52.2% males | Both asymptomatic and symptomatic | Not reported but measured after recovery | Reinfection |
| Cao H 2020 (46) | China | Hospital | Case series | 8 | Mean 54 (26 -72 years) | 37.5% males | Hospitalized and discharged | Day 28 - 50 | Reinfection, Post humoral |
| Breathnach 2021 (47) | UK | Community | Cohort | 66001 | Mean 50 yrs | 60% female | NR | > 7 months | Re -infection and effectiveness |
| An 2020 (48) | China | Hospital | Cohort | 262 | NR | 47.9% males | Mild to moderate,  Discharged | 28 days (14 days isolation and 2 additional weeks post isolation) | Reinfection |
| Ali 2021 (49) | Iraq | Hospital | Case series | 829 | NR | NR | Hospitalized and discharged | 5 months | Reinfection, Post-humoral |
| Adrielle dos Santos 2021 (50) | Brazil | Community | Case series | 33 | Mean 39.2 yrs (SD ±8.53) | 79% female | Symptomatic | 18-134 days between first and second qRT-PCR | Reinfection |
| Abu-Raddad 2020 (51) | Qatar | Community | Case series | 133266 | NR | NR | All hospitalized and discharged Symptomatic - Mild | Median 64.5 (Range 45-129) after negative PCR | Reinfection |
| Abdullah 2020 (52) | Brunei Darussalam | Hospital | Case series | 138 | mean age 41.3 ± 17.0 years) | 59.4% male | All hospitalized and discharged | Day 11 after recovery | Reinfection |
| Abu-Raddad 2021 (53) | Qatar | Community | Cohort | 43044 | Median for females - 35yrs, and for males- 38 yrs | 79% male | All hospitalized and discharged | Median 16.3 weeks, range 0 days - 34.6 weeks | Reinfection |

*Preprints. These four studies were still not published in peer reviewed journals at the time of submission of this review

### **Supplementary Table 2 – Hoy risk of bias**

| STUDY | 1 | 2 | 3 | 4 | 5 | 6 | 7 | 8 | 9 | 10 | OVERALL RISK OF BIAS | ROB Category |
| --- | --- | --- | --- | --- | --- | --- | --- | --- | --- | --- | --- | --- |
| Abdullah 2020 | 0 | 0 | 0 | 0 | 1 | 1 | 1 | 1 | 1 | 1 | 6 | Moderate risk |
| Abu-Raddad 2020 | 0 | 0 | 0 | 0 | 1 | 1 | 1 | 1 | 1 | 1 | 6 | Moderate risk |
| Abu-Raddad 2021 | 0 | 0 | 0 | 0 | 1 | 1 | 1 | 1 | 1 | 1 | 6 | Moderate risk |
| Adrielle dos Santos 2021 | 0 | 0 | 0 | 0 | 1 | 1 | 1 | 1 | 0 | 1 | 5 | High risk |
| Ali 2020 | 0 | 0 | 0 | 0 | 1 | 1 | 1 | 1 | 1 | 1 | 6 | Moderate risk |
| An 2020 | 0 | 0 | 0 | 0 | 1 | 1 | 1 | 1 | 1 | 1 | 6 | Moderate risk |
| Breathnach 2021 | 0 | 0 | 0 | 0 | 1 | 1 | 1 | 1 | 1 | 1 | 6 | Moderate risk |
| Cao H 2020 | 0 | 0 | 0 | 0 | 1 | 1 | 1 | 1 | 1 | 1 | 6 | Moderate risk |
| Cao S 2020 | 1 | 1 | 1 | 1 | 1 | 1 | 1 | 1 | 0 | 1 | 9 | Moderate risk |
| Chen 2020 | 0 | 0 | 0 | 0 | 1 | 1 | 1 | 1 | 1 | 0 | 5 | High risk |
| Chirathaworn 2020 | 0 | 0 | 0 | 0 | 1 | 1 | 1 | 1 | 0 | 1 | 5 | High risk |
| Dan 2021 | 0 | 0 | 0 | 0 | 1 | 1 | 1 | 1 | 1 | 1 | 6 | Moderate risk |
| De Giorgi 2021 | 0 | 0 | 0 | 0 | 1 | 1 | 1 | 1 | 0 | 1 | 5 | High risk |
| Fendler 2020 | 0 | 0 | 0 | 0 | 1 | 1 | 1 | 1 | 1 | 1 | 6 | Moderate risk |
| Fischer 2021 | 0 | 0 | 0 | 0 | 1 | 1 | 1 | 1 | 1 | 1 | 6 | Moderate risk |
| García-Abellán 2021 | 0 | 0 | 0 | 0 | 1 | 1 | 1 | 1 | 1 | 1 | 6 | Moderate risk |
| Grifoni 2021 | 0 | 0 | 0 | 0 | 1 | 1 | 1 | 1 | 1 | 1 | 6 | Moderate risk |
| Gudbjartsson 2020 | 0 | 0 | 0 | 0 | 1 | 1 | 1 | 1 | 1 | 1 | 6 | Moderate risk |
| Hall 2021 | 0 | 0 | 0 | 0 | 1 | 1 | 1 | 1 | 1 | 1 | 5 | Moderate risk |
| Hamed 2020 | 0 | 0 | 0 | 0 | 1 | 1 | 1 | 1 | 1 | 1 | 6 | Moderate risk |
| Hansen B 2021 | 0 | 0 | 0 | 0 | 1 | 1 | 1 | 1 | 1 | 1 | 6 | Moderate risk |
| Hansen H 2021 | 1 | 1 | 1 | 1 | 1 | 1 | 1 | 1 | 1 | 1 | 10 | Low risk |
| Havervall 2020 | 0 | 0 | 0 | 0 | 1 | 1 | 1 | 1 | 1 | 1 | 6 | High risk |
| Huang 2020 | 0 | 0 | 0 | 1 | 1 | 1 | 1 | 1 | 1 | 1 | 7 | Moderate risk |
| Huynh 2021 | 0 | 0 | 0 | 0 | 1 | 1 | 1 | 1 | 1 | 1 | 6 | Moderate risk |
| Jia 2020 | 0 | 0 | 0 | 0 | 1 | 1 | 0 | 1 | 1 | 1 | 5 | High risk |
| Jiang 2020 | 0 | 0 | 0 | 1 | 1 | 1 | 1 | 1 | 1 | 1 | 7 | Moderate risk |
| Ko 2020 | 0 | 0 | 0 | 1 | 1 | 1 | 1 | 1 | 1 | 1 | 7 | Moderate risk |
| Li Y 2020 | 0 | 0 | 0 | 1 | 1 | 1 | 1 | 1 | 1 | 1 | 7 | Moderate risk |
| Liu 2020 | 0 | 0 | 0 | 0 | 1 | 1 | 1 | 1 | 1 | 1 | 5 | Moderate risk |
| Murillo-Zamora 2021 | 0 | 0 | 0 | 1 | 1 | 1 | 1 | 1 | 0 | 1 | 5 | Moderate risk |
| Ogega 2020 | 0 | 0 | 0 | 0 | 1 | 1 | 1 | 1 | 1 | 1 | 6 | Moderate risk |
| Olea 2021 | 0 | 0 | 0 | 1 | 1 | 1 | 1 | 1 | 1 | 1 | 7 | Moderate risk |
| Peng 2021 | 0 | 0 | 0 | 0 | 1 | 1 | 1 | 1 | 1 | 1 | 5 | Moderate risk |
| Perez 2021 | 0 | 1 | 1 | 0 | 1 | 1 | 1 | 1 | 1 | 1 | 8 | Moderate risk |
| Pilz 2021 | 1 | 1 | 1 | 1 | 1 | 1 | 1 | 1 | 1 | 1 | 10 | Low risk |
| Prévost 2020 | 0 | 0 | 0 | 0 | 1 | 1 | 1 | 1 | 1 | 1 | 6 | Moderate risk |
| Qiao 2020 | 0 | 0 | 0 | 1 | 1 | 1 | 1 | 1 | 1 | 1 | 7 | Moderate risk |
| Rydyznski 2020 | 0 | 0 | 0 | 0 | 1 | 1 | 1 | 1 | 1 | 1 | 6 | Moderate risk |
| Sheehan 2021 | 0 | 1 | 0 | 0 | 0 | 1 | 1 | 1 | 1 | 1 | 6 | Moderate risk |
| Shu 2020 | 0 | 0 | 0 | 0 | 1 | 1 | 1 | 1 | 1 | 1 | 6 | Moderate risk |
| Wong 2020 | 0 | 0 | 0 | 1 | 1 | 1 | 1 | 1 | 1 | 1 | 7 | Moderate risk |
| Wu J 2020 | 0 | 0 | 0 | 1 | 1 | 1 | 1 | 1 | 1 | 1 | 6 | Moderate risk |
| Wu X 2020 | 0 | 1 | 0 | 1 | 1 | 1 | 1 | 0 | 0 | 1 | 5 | Moderate risk |
| Xiang 2020 | 0 | 0 | 0 | 0 | 1 | 1 | 1 | 1 | 1 | 1 | 6 | Moderate risk |
| Xiao 2020 | 0 | 0 | 0 | 1 | 1 | 1 | 1 | 1 | 1 | 1 | 7 | Moderate risk |
| Ye 2020 | 0 | 0 | 0 | 1 | 1 | 1 | 1 | 1 | 1 | 1 | 7 | Moderate risk |
| Yuan B 2020 | 0 | 0 | 0 | 0 | 1 | 1 | 1 | 1 | 1 | 1 | 6 | Moderate risk |
| Yuan J 2020 | 0 | 0 | 0 | 1 | 1 | 1 | 1 | 1 | 0 | 1 | 6 | Moderate risk |
| Zhang 2020 | 0 | 0 | 0 | 1 | 1 | 1 | 1 | 1 | 1 | 1 | 7 | Moderate risk |
| Zhao J 2020 | 0 | 0 | 0 | 1 | 1 | 1 | 1 | 1 | 1 | 1 | 7 | Moderate risk |
| Zhao Y 2020 | 0 | 0 | 0 | 0 | 1 | 1 | 1 | 1 | 1 | 1 | 6 | Moderate risk |
| Zheng 2020 | 0 | 0 | 0 | 1 | 1 | 1 | 1 | 1 | 1 | 1 | 7 | Moderate risk |
| Zhu 2020 | 0 | 0 | 0 | 0 | 1 | 0 | 1 | 1 | 1 | 1 | 5 | High risk |

Risk of bias using the tool by Hoy et al 2012. High risk, and unclear scored as zero “0”, while low risk was scored as one “1” for each item. Total score out of 10 where 10 depicts low overall risk and 0 high overall risk. Scores 0-5 (high risk), 6-8 (moderate risk) and 9-10 (low risk)

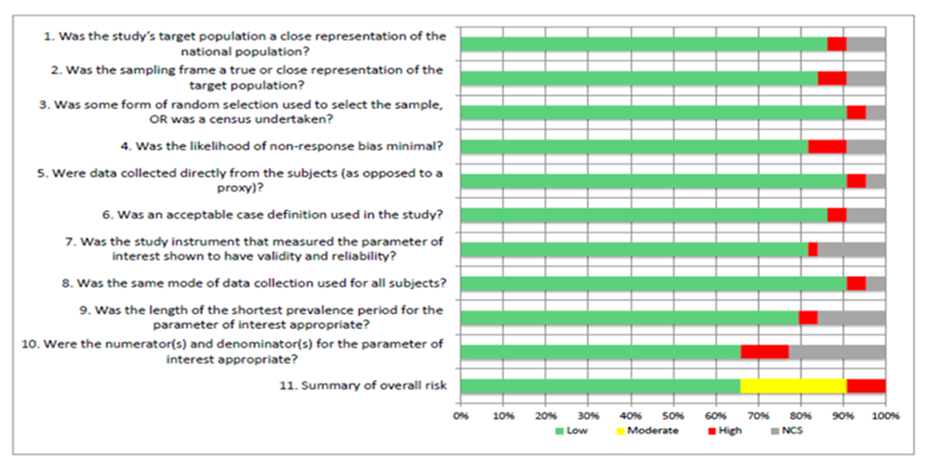

### **Supplementary Table 3 – MASTER assessment for comparative studies**

| Safeguard item | Pilz 2021 | Sheehan 2021 | Hansen 2021 | Breathnach 2021 | Hall 2021 |
| --- | --- | --- | --- | --- | --- |
| 1.    Data collected after the start of the study was not used to exclude participants or to select them into the analysis | 1 | 1 | 1 | 1 | 1 |
| 2.    Participants in all comparison groups met the same eligibility requirements and were from the same population and timeframe | 0 | 0 | 0 | 0 | 0 |
| 3.    Determination of eligibility and assignment to treatment group/exposure strategy was synchronized | 0 | 0 | 0 | 0 | 0 |
| 4. None of the eligibility criteria were common effects of exposure and outcome | 1 | 1 | 1 | 0 | 1 |
| 5.    Any attrition (or exclusions after entry) is less than 20% of total participant numbers | 0 | 0 | 0 | 0 | 0 |
| 6.    Missing data is less than 20% | 0 | 0 | 1 | 0 | 1 |
| 7.    Analysis accounted for missing data | 0 | 0 | 1 | 0 | 1 |
| 8.    Exposure variations/treatment deviations were less than 20% | 0 | 1 | 1 | 1 | 1 |
| 9.    Variations in exposure or withdrawals after the start of the study were addressed by analysis | 0 | 1 | 1 | 1 | 1 |
| 10.  Procedures for data collection of covariates were reliable and the same for all participants | 0 | 0 | 1 | 0 | 0 |
| 11. The outcome was objectively defined and/or reliably measured | 1 | 1 | 1 | 1 | 1 |
| 12. Exposures/ interventions were objectively defined and/ or reliably measured | 1 | 0 | 1 | 0 | 0 |
| 13. Outcome assessor(s) were blinded | 0 | 0 | 0 | 0 | 0 |
| 14. Participants were blinded | 0 | 0 | 0 | 0 | 0 |
| 15. Caregivers were blinded | 0 | 0 | 0 | 0 | 0 |
| 16. Analyst was blinded | 0 | 0 | 0 | 0 | 0 |
| 17. Care was delivered equally to all participants | 0 | 0 | 0 | 0 | 0 |
| 18. Cointerventions that could impact the outcome were comparable between groups or avoided | 0 | 0 | 0 | 0 | 0 |
| 19. Control and active interventions/ exposures are sufficiently distinct | 0 | 0 | 0 | 0 | 0 |
| 20. Exposure/intervention definition consistently applied to all participants | 1 | 1 | 1 | 1 | 0 |
| 21. Outcome definition consistently applied to all participants | 1 | 1 | 1 | 1 | 1 |
| 22. The time period between exposure and outcome is similar across patients and between groups or the analyses adjust for different lengths of follow-up of patients | 1 | 1 | 1 | 1 | 0 |
| 23. Design and/or analytic strategies were in place that addressed potential confounding | 0 | 0 | 1 | 0 | 1 |
| 24. Key confounders addressed through design or analysis were not common effects of exposure and outcome | 0 | 0 | 0 | 0 | 0 |
| 25. Key baseline characteristics / prognostic indicators for the study were comparable across groups | 0 | 1 | 0 | 0 | 0 |
| 26. Participants were randomly allocated to groups with adequate randomisation process | 0 | 0 | 0 | 0 | 0 |
| 27. Allocation procedure was adequately concealed | 0 | 0 | 0 | 0 | 0 |
| 28. Conflict of interests were declared and absent | 1 | 1 | 1 | 1 | 1 |
| 29. Analytic method was justified by study design or data requirements | 0 | 0 | 1 | 0 | 1 |
| 30. Computation errors or contradictions were absent | 1 | 1 | 1 | 1 | 1 |
| 31. There was no discernible data dredging or selective reporting of the outcome | 1 | 1 | 1 | 1 | 1 |
| 32. All subjects were selected prior to intervention/exposure and evaluated prospectively | 0 | 0 | 0 | 0 | 0 |
| 33. Carry-over or refractory effects were avoided or considered in the design of the study or were not relevant | 1 | 1 | 1 | 1 | 1 |
| 34. The intervention/ exposure period was long enough to have influenced the study outcome | 1 | 1 | 1 | 1 | 1 |
| 35. Dose of intervention/ exposure was sufficient to influence the outcome | 1 | 1 | 1 | 1 | 1 |
| 36. Length of follow-up was not too long or too short in relation to the outcome assessment | 1 | 1 | 1 | 1 | 1 |
| Summary count of safeguard items | 14 | 16 | 21 | 14 | 17 |

### **Supplementary Figure 1 – Proportion with detectable IgG after within 1 month after recovery from COVID-19**

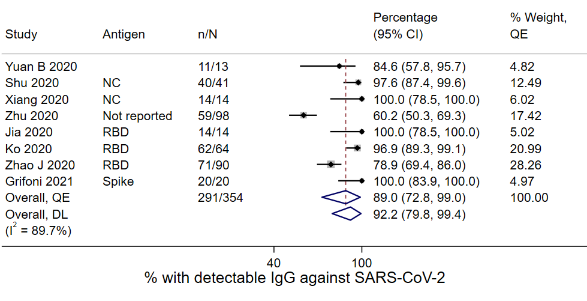

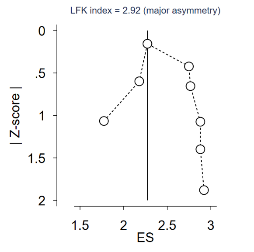

### **Supplementary Figure 2 – Proportion with detectable IgG after 1-<3 months after recovery from COVID-19**

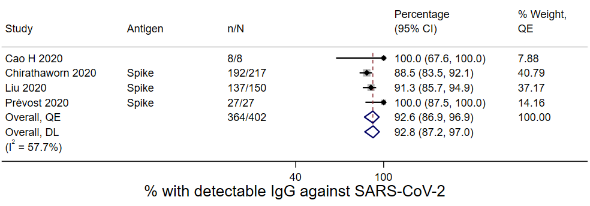

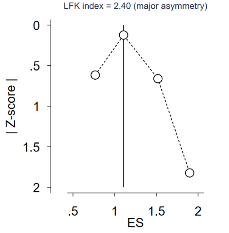

### **Supplementary Figure 3 – Proportion with detectable IgG after 3-<6 months after recovery from COVID-19**

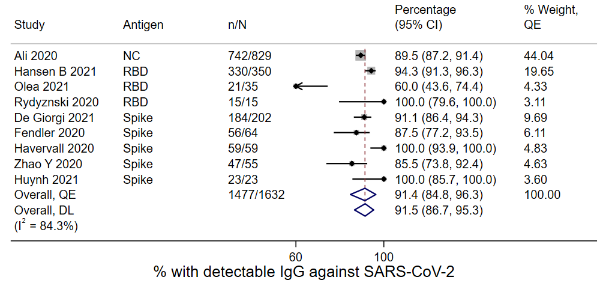

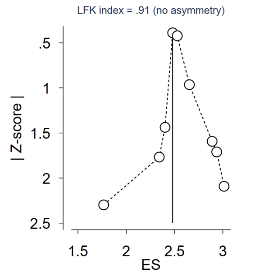

### **Supplementary Figure 4 – Proportion with detectable IgG after ≥6 months after recovery from COVID-19**

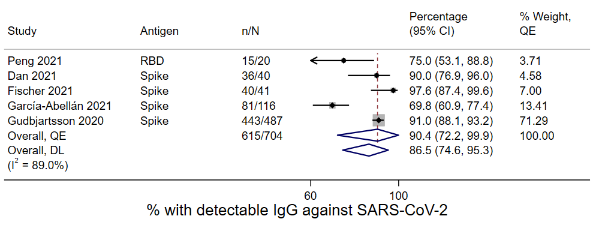

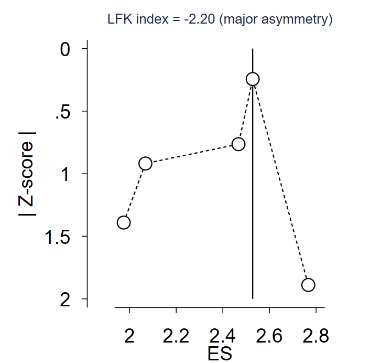

### **Supplementary Figure 5 – Proportion with detectable memory CD4+ T cells after recovery from COVID-19**

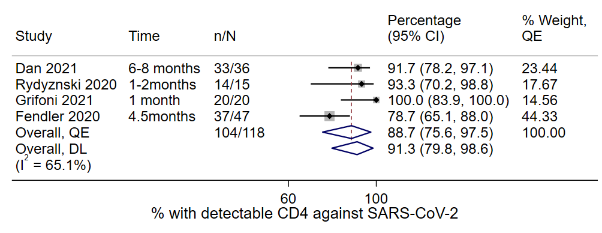

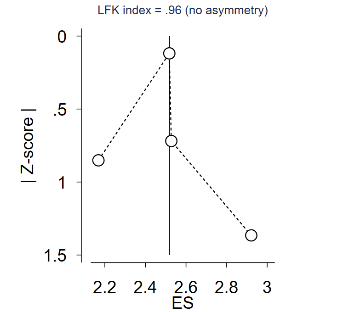

### **Supplementary Figure 6 – Proportion with detectable memory CD8+ T cells after recovery from COVID-19**

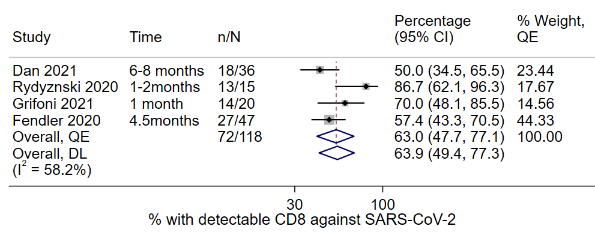

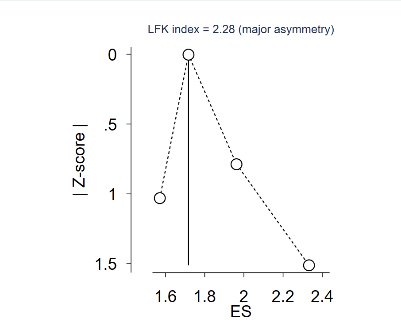

### **Supplementary Figure 7 – Doi plot for proportion with possible reinfection ≥3months after recovery from COVID-19**

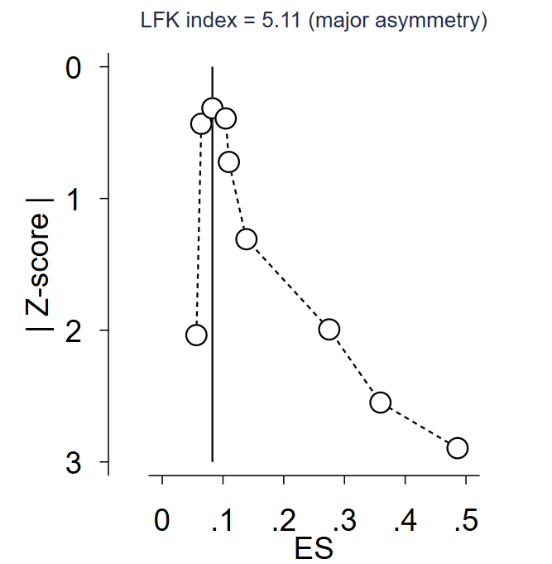

### **Supplementary Figure 8 – Proportion testing positive within 1 month after recovery from COVID-19**

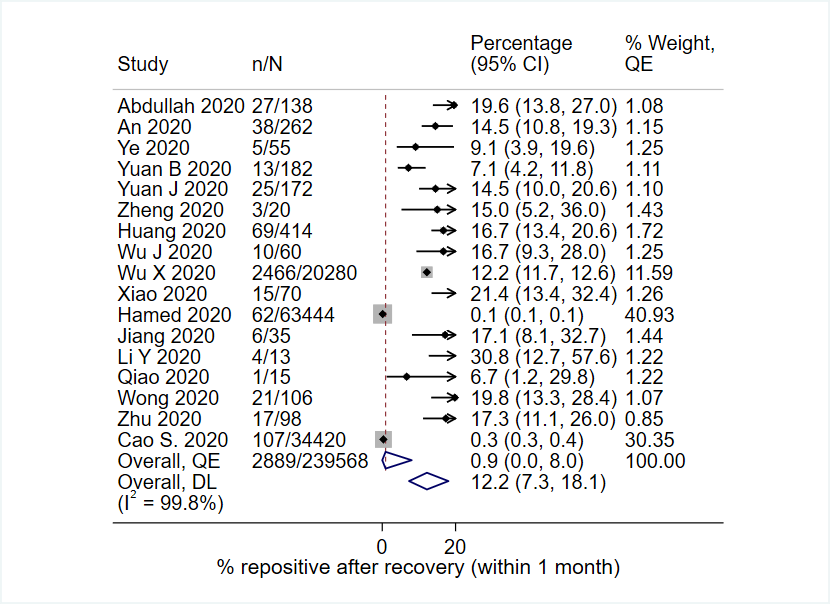

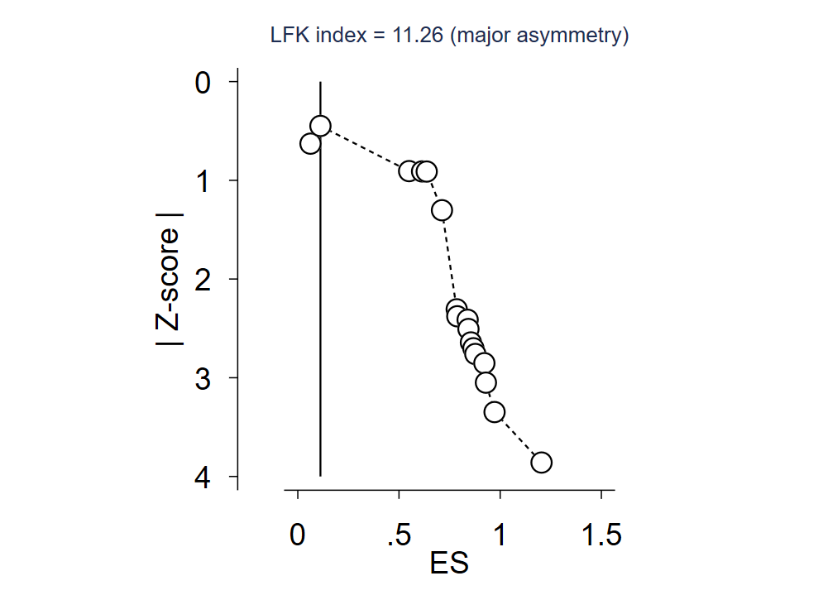

### **Supplementary Figure 9 – Proportion testing positive at 2-3months after recovery from COVID-19**

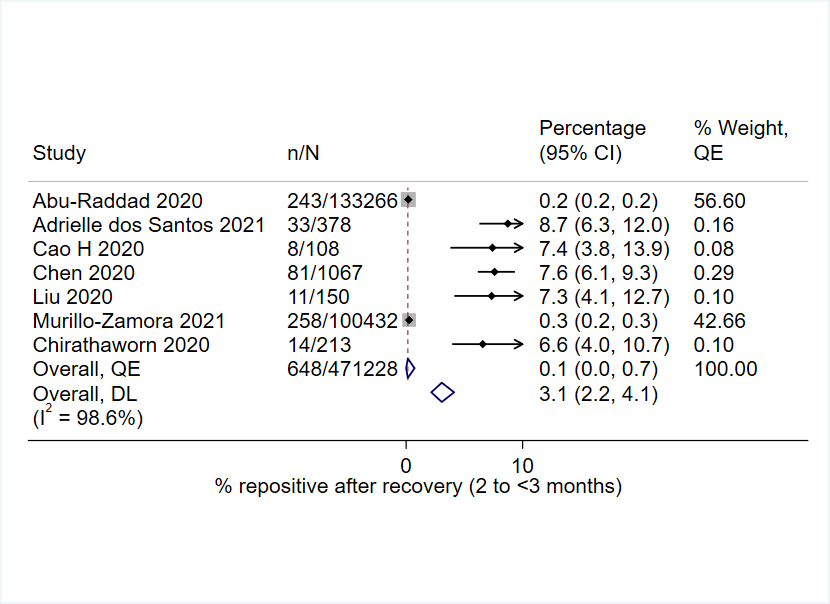

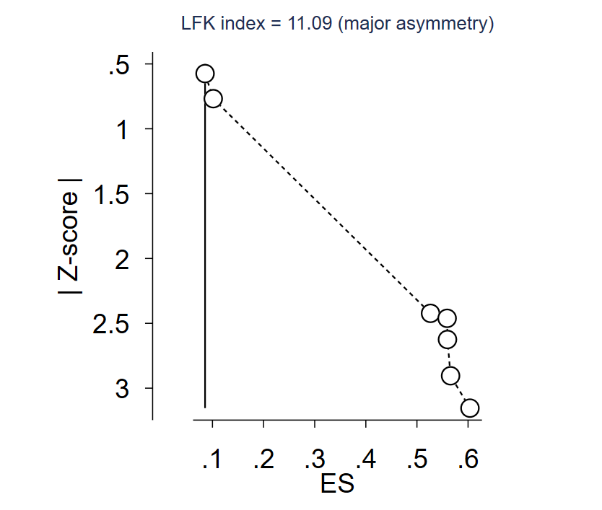

### **Supplementary Figure 10 – Doi plot for proportion for efficacy of previous COVID-19 in preventing future infection**

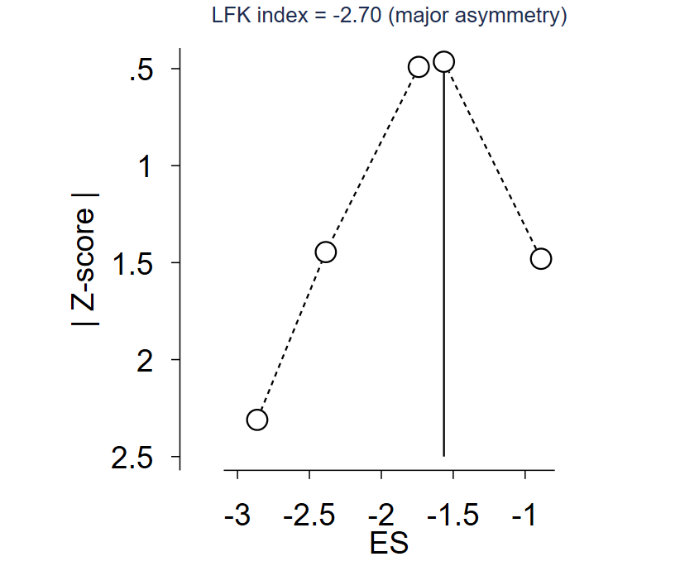

### **Supplementary Table 4 – Findings from previous systematic reviews**

| **Author, year** | **Design** | **N# of studies** | **N# of prtcpnts** | **Longest length of follow up** | **IgG prevalence at follow up** | **IgM** | **T cell**  **CD4** | **T cell**  **CD8** | **Memory B** | **Reinfection / repositive** | **Comments** |
| --- | --- | --- | --- | --- | --- | --- | --- | --- | --- | --- | --- |
| Váncsa (2021) (54) | Systematic review | 56 | 123 | >60 days | IgG positive in 86.1% (n = 31/36) at first episode and  94.2% (n = 49/52) at second episode | NR | NR | NR | NR | 96 (78%) at ≤60 days and 27 (22%) at >60 days | Included case reports. |
| Wu Yan (2021) (55) | Network meta-analysis | 71 | 8, 647 |  | NR | NR | 2216 patients from 14 studies reported  the differences in CD4+ T cells counts | 12 studies involved 2091 patients reported about CD8+ T cells levels | NR | NR | Length of follow up not reported but the reported studies were conducted between January 2020 and March, ranging between 7 days and 2months. |
| Choudhary (2021) (56) | Systematic review | 16 | 20 | 44 days to 282 days | NR | NR | NR | NR | NR | 25% (5/20) at 2 months |  |
| Piri (2021) (57) | Systematic review | 66 | 1128 | 1 – 140 days | Ranged between 58.8 – 100% | Ranged between 11 – 95% | NR | NR | NR | NR |  |
| Arafkas (2021) (58) | Systematic review | 15 including an in vivo study | 215 | 39 ± 9 days | NR | NR | NR | NR | NR | 4% (8/215) at 1 month | Reinfection rate calculated while extracting |
| Farrukh (2020) (59) | Systematic review | 27 | 1616 | >1 month | 166 (10%) at >1 month | 165 (10%) at >1 month | NR | NR | NR | 253 (16%) tested positive the second time after an average  period of 11.5 days from the last negative RT‐PCR test | 1. Data recalculated.  2. article mixing up repositivity and reinfection |
| Bwire (2020) (60) | Systematic review | 6 | 11 | NR | 9/11 (82%) at birth | 8/11 (73%) at birth | NR | NR | NR | NR | Studied children. |
| Shrotri  (2021) (61) | Narrative review |  |  |  |  |  |  |  |  |  |  |
| Iwamura et al (2021) (62) | Narrative review |  |  |  |  |  |  |  |  |  |  |

NR – Not reported
